# Reward network modulation while learning movements to music during weekly multisensory training program for people with Parkinson’s disease: mood effects over 7-yrs

**DOI:** 10.1101/2025.07.19.25331836

**Authors:** Emily Marie D’Alessandro, Ashkan Karimi, Karolina A Bearss, Rachel J Bar, Joseph F.X. DeSouza

**Affiliations:** Department of Psychology, Centre for Vision Research, York University, Toronto, ON, Canada; Department of Psychology, Algoma University, Brampton, ON, Canada; Canada’s National Ballet School, Toronto, ON, Canada; Multisensory Neuroscience Translation Laboratory, Vision: Science to Application -VISTA, Connected Minds for a Healthy, Just Society, Canadian Action and Perception Network (CAPnet), York University, Toronto, ON, Canada

## Abstract

Dance is an art form and simultaneously a form of exercise that provides pleasure to individuals who practise it regularly with subsequent improvements to motor and non-motor symptoms in people with Parkinson’s disease (PD). Recent evidence suggests that dance promotes neuroplasticity and correlates with improvements of depression scores in individuals with PD. However, no neuroplasticity studies using fMRI and EEG have been conducted on people with Parkinson’s disease (PwPD) who attend community dance programs. Thus, we present an observational examination across 7-years that explores the effects of non-motor symptoms of anxiety and depression. The study utilized multimodal recordings from functional MRI (fMRI), electroencephalogram (EEG) and measured affective state using the expanded version of the Positive and Negative Affect Schedule (PANAS-X) questionnaire. fMRI was performed at four time points in the first year (September, December, January, and May) where participants visualized/imagined the choreography they had learned in the dance studio while listening to the recording of the music in the MRI. Resting state EEG was recorded immediately before and after the dance class when the dancers volunteered over the 7 years. In addition, mood was monitored using the PANAS-X in conjunction with the EEG sessions. The PANAS-X was sorted by generalized anxiety disorder (positive and negative affect) symptoms, and major depressive disorder (positive and negative affect) symptoms, along with standard general negative affect (GNA) and general positive affect (GPA). This analysis reveals significant improvements in both symptoms after dance and across time. fMRI results show significant BOLD signal changes in the nucleus accumbens and anterior cingulate cortex, along with trends in other regions in the reward pathway such as the ventral tegmental area and the orbitofrontal cortex. rsEEG results show significant increases in alpha peak values after dance, in both EO and EC conditions. The findings support the idea that dance is a multimodal form of neurorehabilitation with beneficial effects on both motor and non-motor symptoms, including anxiety and depression.

## 1. INTRODUCTION

Parkinson’s disease (PD) is the second most common neurodegenerative disorder, preceded only by Alzheimer’s disease. It is marked by the progressive loss of dopaminergic cells in the substantia nigra pars compacta. PD is typically noted by motor symptoms such as bradykinesia, muscle rigidity, poor posture, and resting tremor, but non-motor symptoms such as loss of smell, autonomic dysfunction, sleep disturbances, and emotional changes are often present and premotor markers of this disorder. Depression is both a premotor marker and NMS of PD (Miller & O’Callaghan, 2015). Depressive symptoms are apparent in 35% (Reijnders et al., 2007) and anxiety disorders are apparent in 31% of individuals with PD (Broen et al., 2016).

There is no cure for PD, however treatments exist to mask symptoms and increase quality of life. Common treatments for PD are levodopa and dopamine agonists, but they are not always effective and have many side effects, such as dyskinesia overtime. Deep brain stimulation (DBS) is another treatment that specifically targets subcortical structures; however, it is highly invasive as electrodes are surgically implanted into the brain (Stoker & Barker, 2020). The administration of antidepressants also has negative side effects. Read et al. (2020) found that 42.7% of antidepressant users reported entirely negative experiences with their medication. These users reported physical side effects, including dry mouth, insomnia, loss of libido, dizziness, headaches, and weight gain. They also experienced withdrawal effects, such as headaches and nausea when attempting to reduce or stop their dosage, along with emotional and cognitive numbing. They also reported altered prescriber relationships, which included a lack of information about side effects, other treatments, and support on behalf of their physician. Benzodiazepines are common prescriptions for anxiety, but have adverse side effects including drowsiness, psychomotor slowing, decreased IQ, and delayed response time, among others (Stewart, 2005).

Dance is beneficial for the elderly population as it is a multimodal form of physical activity that engages learning, attention, memory, emotion, motor coordination, balance, gait, visuospatial abilities, imagination, and social interaction (Kshtriya et al., 2015). Dance has shown to be an effective additive treatment method for people with PD (PwPD) by putatively promoting neuroplasticity in regions such as the supplementary motor area (SMA), pre-SMA, auditory cortex, insula, and temporal gyri and improving motor and NMS (DeSouza et al., 2023; Simon et al., 2024; Barnstaple et al., 2022; Rabinovich et al., 2021, Bearss et al., 2017). An increasing body of research over the past 20 years has demonstrated that dance can be a beneficial treatment for Parkinson’s disease (PD), helping to reduce motor symptoms by improving balance and decreasing symptom severity, while also enhancing non-motor symptoms such as mood (Fontanesi & DeSouza, 2021; Westheimer et al., 2015; Houston & McGill, 2013; Garlovsky, Overton & Simpson, 2016).

Bearss and DeSouza (2021) were the first study that found no Parkinson’s disease progression of both PD motor symptoms and NMS symptoms over three years of weekly community dancing. This study proposes that dance could lead to neuroprotective effects in brain networks described in the case study of Simon et al 2024. Asmundson et al. (2013) found that exercise programs lasting 16 weeks or more have a protective effect against the development of anxiety in older adults which could be helpful for reducing anxiety in our PwPD. In addition, PwPD have shown lower electroencephalogram (EEG) complexity in gamma, beta, and alpha waves (Yi et al., 2016) and oscillatory resting slowing with cognitive decline and motor deterioration (Olde Dubbelink et al., 2013). However, there is yet to be a dance-related study that evaluates both anxiety and depression-related symptoms and correlates these to both blood-oxygenation level dependent (BOLD) signal changes and resting state electroencephalogram (rsEEG).

The present study reports on an observational longitudinal investigation over 7-years of the effects of dance training on depression and anxiety-related symptoms in PwPD and control participants. PANAS-X surveys and rsEEG data were collected before and after dance classes from 2014-2019, to examine changes in affective state on one to two subjects each week. Functional magnetic resonance (fMRI) was completed at four time points across an 8-month training period to examine changes in cortical activity while participants imagined the learned dance. We hypothesize that dance will improve depression and anxiety-related symptoms and these will be followed by modulation of brain signals in the reward pathway. Regions of interest (ROIs) related to depression are regions in the reward pathway such as the orbitofrontal cortex (OFC), ventral tegmental area (VTA), and nucleus accumbens (NAc), along with the insula. ROIs related to anxiety are the prefrontal cortex (PFC), anterior cingulate cortex (ACC), and posterior cingulate cortex (PCC) based on Mochcovitch et al. (2014) and Zhao et al. (2006) studies. We anticipate significant changes in BOLD signals in those listed regions. We hypothesize a change in the alpha wave signal after dance, due to its role in attention and arousal, relating to changes in depression and anxiety-related symptoms (Gutmann et al., 2015). We predict that the statistical analysis of the PANAS-X surveys will show a positive correlation with changes in the BOLD signal and brain oscillations in the resting-state EEG (rsEEG) data.

Su et al. (2022) observed alterations in the posterior cingulate gyrus, supplementary motor area (SMA), and cerebellum during the resting state. This demonstrates that the SMA may have more involvement in affective state than initially thought. There is also evidence of alterations in the mesolimbic and mesocortical pathways in individuals with Parkinson’s disease (PwPD) and depression (Wei et al., 2018). This supports the hypothesis and previous evidence of the involvement of the reward pathway in depression. With this, paired with alterations in the basal ganglia network, we expect to find significant changes in the VTA (ventral tegmental area), along with other regions of the mesolimbic pathway.

According to a systematic review by Carey et al. (2020), regions that exhibited significant changes in the BOLD signal in PwPD related to anxiety, which align with our regions of interest (ROIs), include the anterior cingulate cortex (ACC) and strong functional connectivity between the striatum and the temporal cortex, as well as between the striatum and the cingulate cortex. This evidence supports the association between Parkinson’s disease and anxiety, highlighting the involvement of the striatum.

## 2. METHODS

### 2.1 Dance training

Participants attended weekly Dance for PD classes at Canada’s National Ballet School (NBS) and Trinity St. Paul’s Church Studio in Toronto, Ontario. The dance classes lasted 75 minutes and incorporated elements of ballet, jazz, Argentinian tango, dance theatre, freestyle, and choreographed movements, all taught by a certified Dance for Parkinson’s Disease (Dance for PD) instructor to live piano music. Each class began with an opening, introducing themselves accompanied by a dance movement. The warm-up included stretching their distal joints and limbs. The class began seated, leading to paired mirror dance with a partner, and balance practice. Finally, participants began to dance to the triplet rhythm of waltz by starting with a seated dance shuffle to gradually increase the amount of dance movements, leading up to a 2-minute performance facing their partner with steps and movement sequences (Bearss & DeSouza 2021; Simon et al. 2024).

### 2.2 rsEEG procedure

rsEEG was measuring putative neuroplastic changes in brain activity associated with participating in one dance class. The recording included 3 minutes of eyes closed (EC) and 3 minutes of eyes open (EO) at random, using a 14-channel EPOC X EEG neuroheadset. To be included in the neuroimaging analysis the subject had to be in both fMRI and an EEG imaging session. rsEEG data of six participants were analyzed, all of whom volunteered in the fMRI part, and 5 of whom also participated in PANAS-X. Participants were between the ages of 61-72 (Mean Age = 68; SD = 4.183300133; 2 females, 4 males). 1 of the 6 participants’ age was not reported.

### 2.3 Scanning procedure

10 participants with PD were scanned for a minimum of 1-4 times throughout the 8- month dance period between the ages of 60-75 (Mean Age = 67.85714286; SD = 4.84522347). 3 ages were not reported. 9 participants were recorded at the first scan (T1) in September. 7 participants were scanned for the second time (T2) in December. 7 participants were scanned for the third time (T3) in January. 5 participants were scanned for the fourth time (T4) in May. The participants were scanned at the Sherman Health Science Research Centre at York University. Consent and safety screening and task training was provided before each scanning session.

The anatomical and functional scans were captured using a 32-channel head coil in a 3T Siemens Tim Trio MRI scanner. A total of 28 scans were acquired. Each scan was 240 volumes. The participants were instructed to keep their eyes open or closed during visualization in the scanner and their head was secured in place to prevent movement artifacts. All participants reported that they closed their eyes during the task visualization. The participants visualized the learned dance choreography from the studio while in the scanner, listening to music through headphones during the visualization. The experimental design utilized a block format consisting of a 60-second ON state (dance visualization accompanied by music) followed by a 30-second OFF state (fixation block with no music). The sequence began with the fixation block and alternated with the visualization block, repeated a total of five times, resulting in a total protocol duration of 8 minutes. The ON state was cued to music recorded in the National Ballet School (NBS) studio from a dance class conducted in September.

### 2.4 PANAS-X statistical analysis

The PANAS-X is a survey measuring positive and negative affect by rating a set of 60 affective states on a scale from 1 to 5 (past two weeks). Participants completed the survey before and after each dance class, indicating the extent to which they had felt this way before each class, followed by their feelings after the dance class. Ratings varied from 1 - very slightly or not at all; 2 - a little; 3 - moderately; 4 - quite a bit and 5 - extremely. The PANAS-X is normally divided into general dimension scales (negative affect and positive affect), basic negative emotion scales (fear, hostility, guilt, and sadness), basic positive emotion scales (joviality, self-assurance, and attentiveness) and other affective states (shyness, fatigue, serenity and surprise) and each of these categories have 3-10 affective items of the 60-item list (Watson & Clark, 1994).

We chose to parse the PANAS-X into a depression and anxiety measure. By comparing the item composition of the PANAS-X scales to the diagnostic criteria in the DSM-V for Major Depressive Disorder (MDD) and Generalized Anxiety Disorder (GAD), the affective states used for depression and anxiety-related symptoms are shown in Table 1.

**Table 1:** Displays the categorization of the depression-related symptoms, anxiety-related symptoms, general negative affect (GNA) and general positive affect (GPA) statistical analysis based on the item composition of the PANAS-X scales.

|  |  |
| --- | --- |
| **Depression negative affect** | *Guilt (6): guilty, ashamed, blameworthy, angry at self, disgusted with self, dissatisfied with self*<br>*Sadness (5): sad, blue, downhearted, alone, lonely*<br>*Fatigue (4): sleepy, tired, sluggish, drowsy* |
| **Depression positive affect** | *Joviality (8): cheerful, happy, joyful, delighted, enthusiastic, excited, lively, energetic*<br>*Self-assurance (6): proud, strong, confident, bold, fearless, daring*<br>*Attentiveness (4): alert, attentive, concentrating, determined* |
| **Anxiety negative affect** | *Fear (6): afraid, scared, frightened, nervous, jittery, shaky*<br>*Hostility (6): angry, hostile, irritable, scornful, disgusted, loathing*<br>*Shyness (4): shy, bashful, sheepish, timid*<br>*Fatigue (4): sleepy, tired, sluggish, drowsy* |
| **Anxiety positive affect** | *Attentiveness (4): alert, attentive, concentrating, determined* |
|  | *Serenity (3): calm, relaxed, at ease* |
| **General Negative Affect (GNA)** | *Negative Affect (10): afraid, scared, nervous, jittery, irritable, hostile, guilty, ashamed, upset, distressed* |
| **General Positive Affect (GPA)** | *Positive Affect (10): active, alert, attentive, determined, enthusiastic, excited, inspired, interested, proud, strong* |

The statistical analysis was conducted using RStudio Version 2023.06.1+524. A two-way ANOVA was used to compare the groups; controls and PwPD, and timing; pre-dance sessions and post-dance sessions. There was a total of 56 participants (n_PD_=37, n_control_=19) with 94 individual PD sessions and 43 individual control sessions. Participants with Parkinson’s disease (PwPD; n_PD_=37) ranged in age from 52 to 87 years (mean = 70.45, SD = 7.95) with controls ranging in age from 22 - 80 (mean = 65.85, SD = 10.81). Five participants’ ages were not reported. The current MRI study is exploratory in nature and does not utilize a randomized controlled design since all volunteers were from the one dance class; therefore, we will not correct for multiple comparisons. The study was designed as a learning paradigm for pre-SMA activation (Bar & DeSouza, 2016; Simon et al 2024; Bearss et al 2024), and modulations in other regions are observational.

### 2.5 Data Processing and Analysis

Pre-processing steps were applied to all rsEEG recordings in MATLAB_R2023b using FieldTrip version 20231025. This sorted the EO and EC data, completed an Independent Component Analysis (ICA), and completed visual inspection of the topographical disposition and data checking. A power spectrum analysis, alpha peak across subjects, and grand average power spectrum was completed to visualize the oscillation changes (Di Nota et al., 2017). Two scans were excluded for failing to run through pre-processing steps.

Pre-processing steps were applied to all fMRI scans to reduce noise, correct for motion and spatial and temporal smoothing. The analysis was performed using BrainVoyager QX (Version 22.0). Regions of interest (ROIs) were Bonferroni-corrected (p < 0.0001) using the General Linear Model: Multi-subject to compare the dance visualization block to the fixation block. ROIs were manually created based on these published studies (Mamiya et al., 2020; Ding et al., 2022; Peterson et al., 2017; Allen et al., 2016; Gong et al., 2022; Sundermann, Olde lütke Beverborg & Pfleiderer, 2014; Zhao et al., 2007). MATLAB_R2023b was used for BOLD signal number transformations. R Studio was used for plotting graphs and performing post-hoc tests.

The current study is exploratory in nature and observational from a community dance class and was not a randomized controlled design and thus we will not correct for multiple comparisons. The study was designed for pre-SMA and auditory cortex stimulation to examine learning related changes in BOLD activation (Bar & DeSouza, 2016; Simon et al., 2023 PsychAriv- submitted to Frontiers).

## 3. RESULTS

The ROIs for depression and anxiety which were significantly activated at a statistical threshold of p < 0.0001 (Bonferroni-corrected) were the for depression: VTA, right and left NAc, and right and left OFC and for anxiety: right and left ACC, and right PCC (specifically, the RSC). Talairach coordinates for all ROIs are provided in the Supplementary material (Table S1).

As shown in **Fig. 1 (A-J)**, there is a change in the reward pathway in the PwPD’s ROIs. In **Fig. 1A**, there was a decrease trend in activation followed by an increase in activation from T2 to T3 (p = 0.09762664) in the VTA. In **Fig. 1C** and **1E**, both right and left NAc demonstrate a decrease in activation over time, with the left NAc showing a trend between T1 and T3 (p = 0.05615957), and T1 and T4 (p = 0.05903631) and is particularly significant in the right NAc from T1 to T3 (p = 0.04027025). In **Fig. 1G**, there is an increase followed by decrease in BOLD signal in the right OFC (rOFC), whereas in **Fig. 1I** the left there is simply a decrease in BOLD signal over time. The rOFC demonstrates a strong trend (p = 0.05787339), along with a trend on the left side (p = 0.07063665). Overall, t-tests (Bonferroni-corrected) performed for the depression ROIs show a trend (p < 0.08) in the reward pathway.

**Figure 1.**
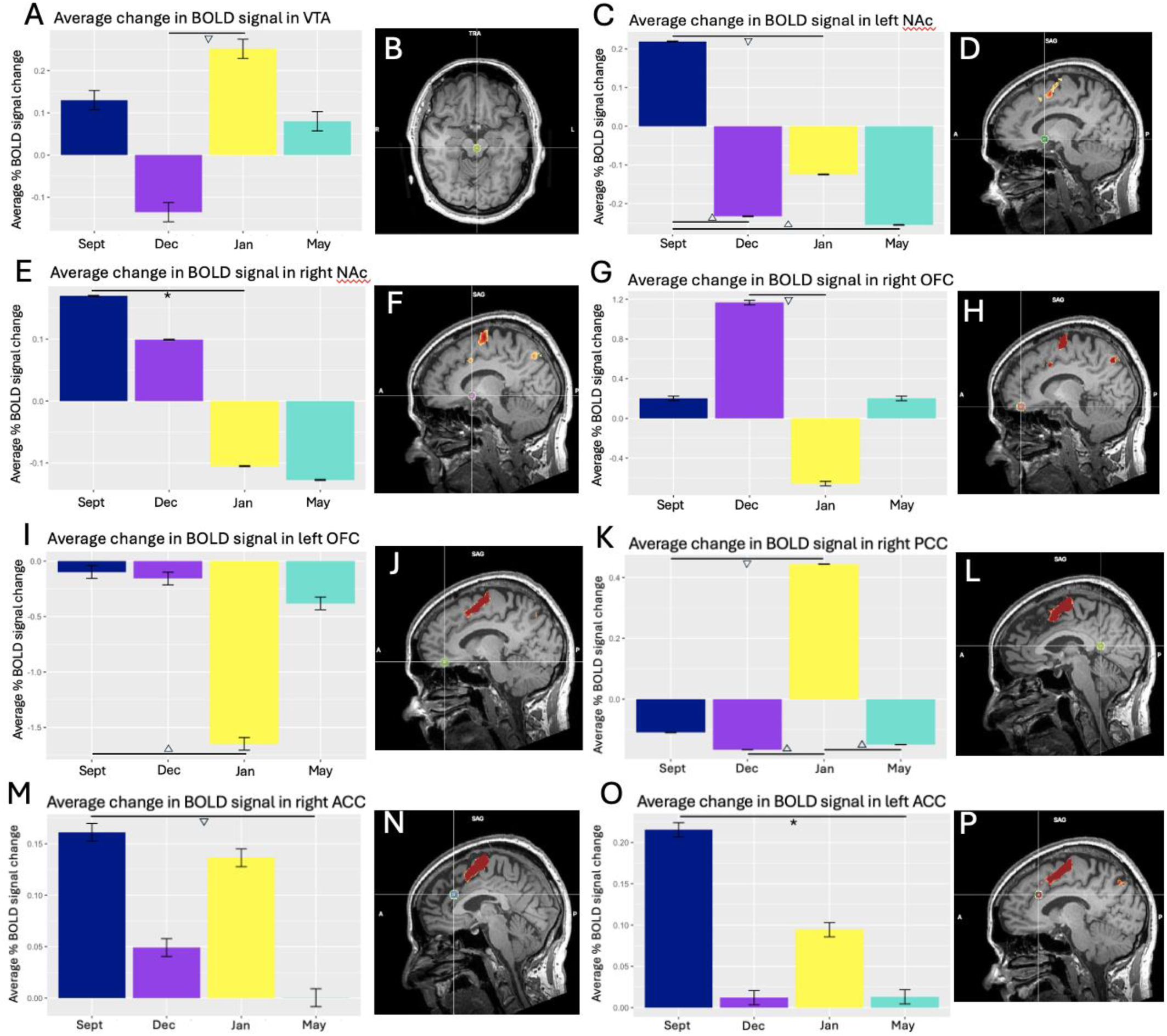
ROIs investigated for depression and anxiety-related changes. **(A)** fMRI average percent BOLD signal change at each time point in the VTA. Error bars represent 0.1 BOLD signal change. A trend is observed from Dec to Jan with p < 0.1 (p=0.098). **(B)** BOLD activation of VTA during dance visualization shown in a horizontal plane, displayed at a statistical threshold of p < 0.0001 (Bonferroni-corrected). **(C)** fMRI average percent BOLD signal change at each time point. Error bars represent 0.1 BOLD signal change. A significant difference is observed between Sept and Jan (p = 0.05615957), between Sept and May (p = 0.05903631), and a trend is seen between Sept and Dec (p = 0.08223606). **(D)** BOLD activation of left NAc (lNAc) during dance visualization shown in sagittal plane, displayed at a statistical threshold of p < 0.0001 (Bonferroni-corrected). **(E)** fMRI average percent BOLD signal change at each time point. Error bars represent 0.10 BOLD signal change. A significant difference is observed between Sept and Jan with p < 0.05 (p = 0.04027025). **(F)** BOLD activation of right NAc (rNAc) during dance visualization shown in sagittal plane, displayed at a statistical threshold of p < 0.0001 (Bonferroni-corrected). **(G)** fMRI average percent BOLD signal change at each time point. Error bars represent 0.1 BOLD signal change. A trend is observed between Dec and Jan with p < 0.08 (p = 0.05787339). **(H)** BOLD activation of right OFC (rOFC) during dance visualization shown in sagittal plane, displayed at a statistical threshold of p < 0.0001 (Bonferroni-corrected). **(I)** fMRI average percent BOLD signal change at each time point. Error bars represent 0.1 BOLD signal change. A trend is observed between Sept and Jan with p < 0.08 (p = 0.07063665). **(J)** BOLD activation of left OFC (lOFC) during dance visualization shown in sagittal plane, displayed at a statistical threshold of p < 0.0001 (Bonferroni-corrected). **(K)** fMRI average percent BOLD signal change at each time point. Error bars represent 0.1 BOLD signal change. A trend is observed between Dec and Jan with p < 0.08 (p = 0.07453964), Jan and May (p = 0.07225377) and Sept and Jan (p = 0.079812). **(L)** BOLD activation of right PCC (specifically the RSC) during dance visualization shown in sagittal plane, displayed at a statistical threshold of p < 0.0001 (Bonferroni-corrected). **(M)** fMRI average percent BOLD signal change at each time point. Error bars represent 0.1 BOLD signal change. A trend is observed between Sept and May with p < 0.10 (p = 0.09802739). **(N)** BOLD activation of right ACC (rACC) during dance visualization shown in sagittal plane, displayed at a statistical threshold of p < 0.0001 (Bonferroni-corrected). **(O)** fMRI average percent BOLD signal change at each time point. Error bars represent 0.1 BOLD signal change. A significant difference is observed between Sept and May with p < 0.05 (p = 0.03883743). **(P)** BOLD activation of left ACC (lACC) during dance visualization shown in sagittal plane, displayed at a statistical threshold of p < 0.0001 (Bonferroni-corrected).

As shown in **Fig. 1 (K-P),** there are trends in the default mode network in the ACC and PCC (p < 0.08). **Fig. 1O** shows the peaks of the modulation in that region. The left ACC (lACC) is particularly interesting with a decrease in BOLD signal across sessions (p = 0.03883743). The retrosplenial cortex (RSC) residing within the right PCC shows a peak in activity at T3, as seen in **Fig. 1K**. This ROI demonstrates a strong trend between T2 and T3 with p < 0.08 (p = 0.07453964), T3 and T4 (p = 0.07225377) and T1 and T3 (p = 0.079812).

The PANAS-X demonstrates statistically significant effects for general negative affect (GNA), general positive affect (GPA), anxiety positive affect, anxiety negative affect, depression negative affect, and depression positive affect scores. The graphs depict healthy controls (HC) and PD before and after dance. Each dot represents a recording session value in the longitudinal graph (each participant completed the survey at least once each). In the first session graph, each dot represents a unique participant. The larger dots extending past the error bars are outliers. Error bars represent a 95% confidence interval (CI). A two-way ANOVA for GNA revealed significant change (Pr(>F)_group_ = 0.00683; Pr(>F)_timing_ = 2.55e-10) in **Fig. 2A** and significant change (Pr(>F)_group_ = 0.00013) in **Fig. 2B**. Post-hoc t-tests (Bonferroni-corrected) revealed significant differences (p < 0.05) between PD-pre and PD-post, PD-pre and HC-pre, PD-pre and HC-post, HC-pre and HC-post, and PD-post and HC-post in **Fig. 2A** and PD-pre and PD-post, PD-pre and HC-post, HC-pre and HC-post, and PD-post and HC-post in **Fig. 2B**.

**Figure 2.**
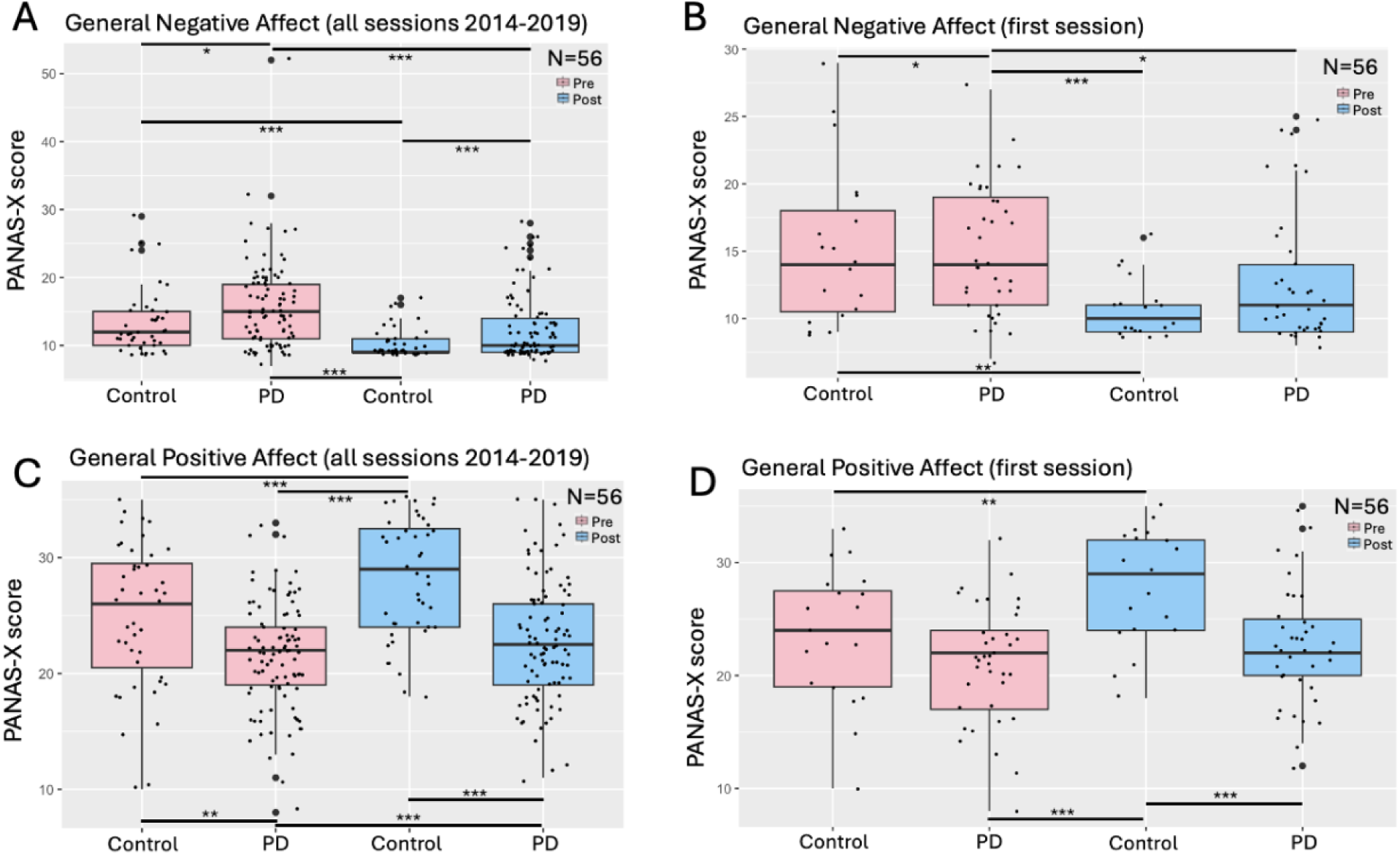
General negative affect and general positive affect scores. **(A)** All recorded sessions from 2014-2019 are plotted. Each dot represents a survey from a volunteer (control/PD). **(B)** Each participant’s first PANAS-X session was extracted from the database and plotted. Each dot represents a participant (n=56). **(C)** All recorded sessions from 2014-2019 are plotted. Each dot represents a survey from a volunteer (control/PD). **(D)** Each participant’s first PANAS-X session was extracted from the database and plotted. Each dot represents a participant (n=56).

A two-way ANOVA for GPA revealed significant change (Pr(>F)_group_ = 1.66e-06; Pr(>F)_timing_ = 2.2e-07, Pr(>F)_group:timing_ = 0.0258) in **Fig. 2C** and significant change (Pr(>F)_group_ = 0.00519; Pr(>F)_timing_ = 0.000265, Pr(>F)_group:timing_ = 0.031313) in **Fig. 2D**. Post-hoc t-tests (Bonferroni-corrected) revealed significant differences (p < 0.05) between PD-pre and PD-post, PD-pre and HC-pre, PD-pre and HC-post, HC-pre and HC-post, HC-pre and PD-post, and PD-post and HC- post in **Fig. 2C** and PD-pre and PD-post, PD-pre and HC-post, HC-pre and HC-post, and PD- post and HC-post in **Fig. 2D**. This demonstrates that dance increases feelings of positive affect.

A two-way ANOVA for anxiety negative affect revealed significant change (Pr(>F)_group_ = 0.000436; Pr(>F)_timing_ = 0.0186) in **Fig. 3A**. Post-hoc t-tests (Bonferroni-corrected) revealed significant differences (p < 0.05) between PD-pre and HC-pre, PD-pre and HC-post, HC-pre and HC-post and PD-post and HC-post in **Fig. 3A** and PD-pre and HC-post and PD-post and HC- post in **Fig. 3B**.

**Figure 3.**
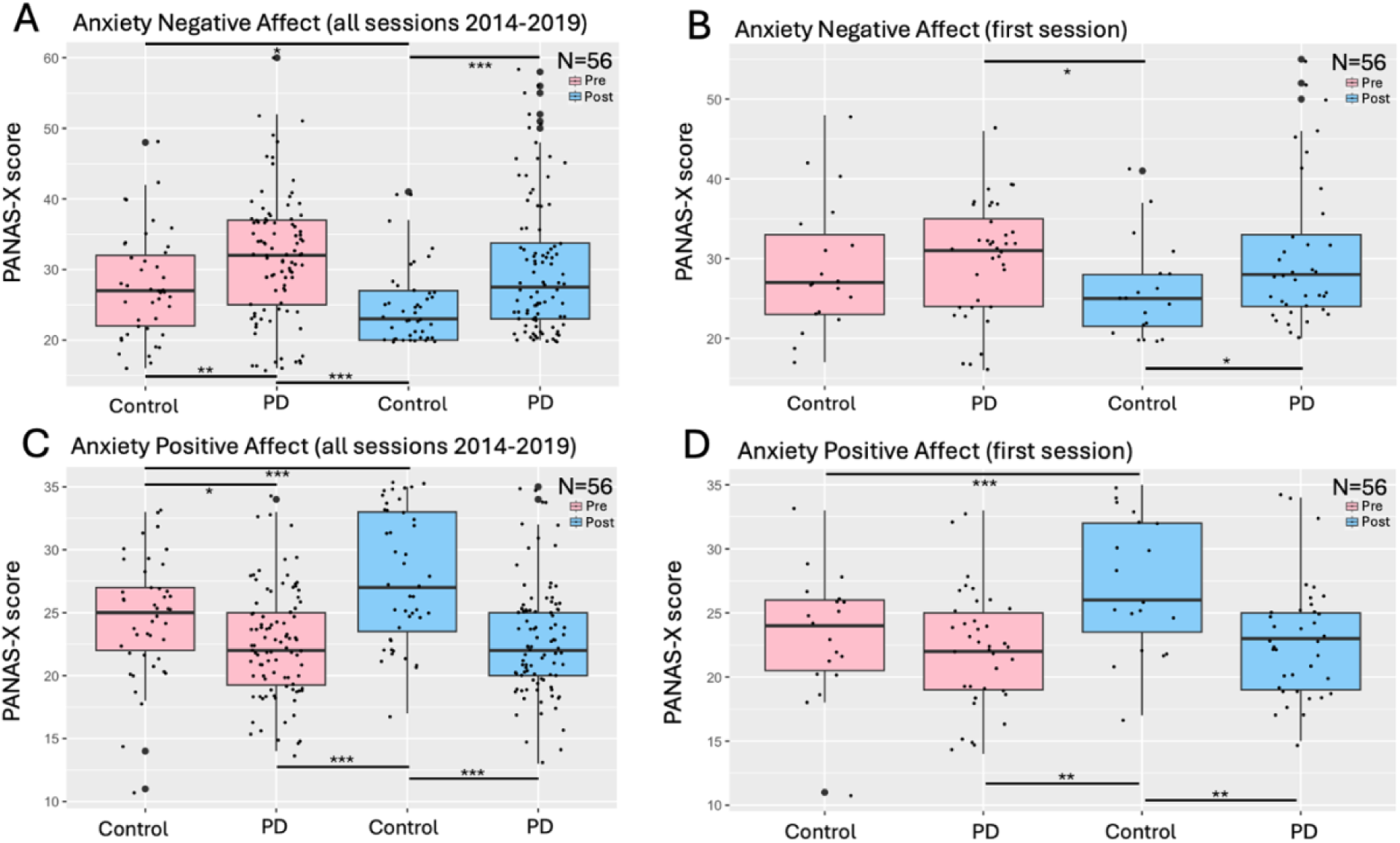
Anxiety negative affect and anxiety positive affect scores. **(A)** All recorded sessions from 2014-2019 are plotted. Each dot represents a survey from a volunteer (control/PD). **(B)** Each participant’s first PANAS-X session was extracted from the database and plotted. Each dot represents a participant (n=56). **(C)** All recorded sessions from 2014-2019 are plotted. Each dot represents a survey from a volunteer (control/PD). **(D)** Each participant’s first PANAS-X session was extracted from the database and plotted. Each dot represents a participant (n=56).

A two-way ANOVA for anxiety positive affect revealed significant change (Pr(>F)_group_ = 1.46e-05; Pr(>F)_timing_ = 0.000216, Pr(>F)_group:timing_ = 0.000118) in **Fig. 3C** and significant change (Pr(>F)_group_ = 0.0244; Pr(>F)_timing_ = 0.00138, Pr(>F)_group:timing_ = 0.00408) in **Fig. 3D**. Post-hoc t-tests (Bonferroni-corrected) revealed significant differences (p < 0.05) between PD-pre and HC- pre, PD-pre and HC-post, HC-pre and HC-post, and PD-post and HC-post in **Fig. 3C** and PD-pre and HC-post, HC-pre and HC-post, and PD-post and HC-post in **Fig. 3D**.

A two-way ANOVA for depression negative affect revealed significant change (Pr(>F)_group_ = 0.00565; Pr(>F)_timing_ = 1.88e-15) in **Fig. 4A** and significant change (Pr(>F)_timing_ = 1.73e-06) in **Fig. 4B**. Post-hoc t-tests (Bonferroni-corrected) revealed significant differences (p < 0.05) between PD-pre and PD-post, PD-pre and HC-pre, PD-pre and HC-post, HC-pre and PD-post, HC-pre and HC-post, and PD-post and HC-post in **Fig. 4A** and PD-pre and PD-post, PD-pre and HC- post, HC-pre and PD-post, HC-pre and HC-post, and PD-post and HC-post in **Fig. 4B**.

**Figure 4.**
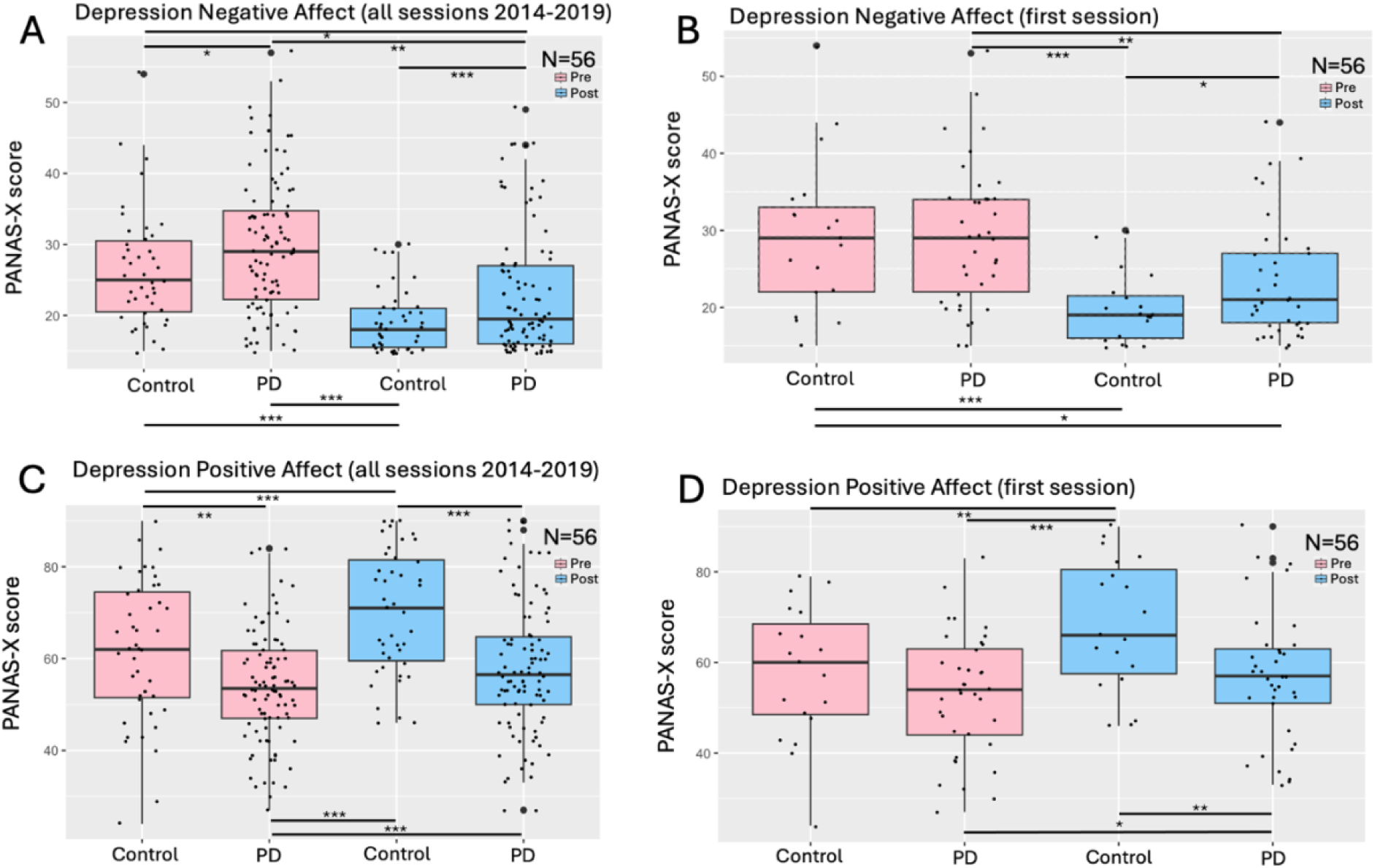
Depression negative affect and depression positive affect scores. **(A)** All recorded sessions from 2014-2019 are plotted. Each dot represents a survey from a volunteer (control/PD). **(B)** Each participant’s first PANAS-X session was extracted from the database and plotted. Each dot represents a participant (n=56). **(C)** All recorded sessions from 2014-2019 are plotted. Each dot represents a survey. **(D)** Each participant’s first PANAS-X session was extracted from the database and plotted. Each dot represents a volunteer (n=56) from a volunteer (control/PD).

A two-way ANOVA for depression positive affect revealed significant change (Pr(>F)_group_ = 2.48ee-05; Pr(>F)_timing_ = 4.87e-08; Pr(>F)_group:timing_ = 0.0237) in **Fig. 4C** and significant change (Pr(>F)_group_ = 0.0285; Pr(>F)_timing_ = 0.000194; Pr(>F)_group:timing_ = 0.037132) in **Fig. 4D**. Post-hoc t-tests (Bonferroni-corrected) revealed significant differences (p < 0.05) between PD-pre and PD-post, PD-pre and HC-pre, PD-pre and HC-post, HC-pre and HC-post, and PD-post and HC-post in **Fig. 4C** and PD-pre and PD-post, PD-pre and HC-post, HC-pre and HC-post, and PD-post and HC-post in **Fig. 4D**. Specific p-values are reported in the Supplemental materials (S1 - S7).

Linear regressions were performed between the average change in PANAS-X scores (positive affect, negative affect, depression negative affect and anxiety negative affect) with average change in BOLD signal across sessions in each ROI (VTA, rNAc, lNAc, rOFC, lOFC, rACC, lACC, rPCC).

Average change in BOLD signal was found by subtracting sessions and then averaging the differences between sessions (i.e. (T4-T3) + (T3-T2) + (T2-1) / 3). For PANAS-X values, for participants that completed more than one session, their sessions were averaged, followed by taking the difference between the pre- and post-average session scores.

As shown in **Fig. 5**, one significant regression was found in the rPCC (p = 0.05754; R^2^ = 0.7503; Adjusted R^2^ = 0.6671; F-statistic = 9.017). For rsEEG alpha peak values, and PANAS-X scores (positive affect, negative affect, depression negative affect, depression positive affect, anxiety negative affect and anxiety positive affect) no significant correlations were determined. Changes in peak values were determined by subtracting pre and post condition peak values, in EO and EC conditions respectively.

**Figure 5.**
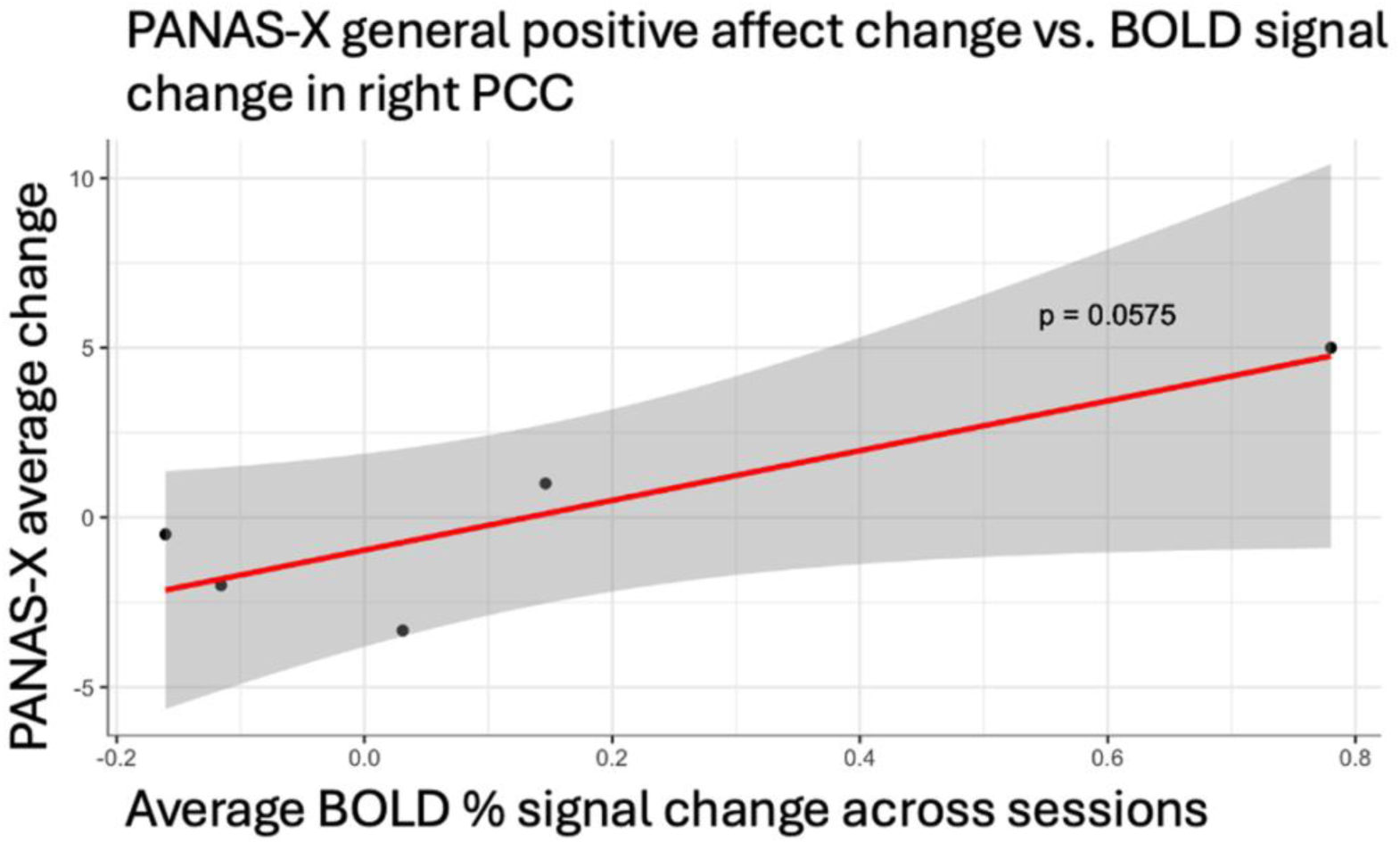
Average BOLD % signal change is plotted on the x-axis with PANAS-X average change in general positive affect scores plotted on the y-axis. There is a trend demonstrating an increase in PANAS-X scores as the average BOLD % signal increases.

Analyses for various conditions were performed for rsEEG EO and EC conditions. Figures of the heads display the global alpha wave activation across the 14 channels. The dots on the diagram represent each channel. Boxplots were plotted to perform an analysis between the pre and post-dance alpha frequency peak power values and pre- and post-dance alpha peak values and a paired t-test (Bonferroni-corrected) was performed.

All participants’ scans (n=6, sessions=20) are plotted, along with boxplots for the alpha frequency and alpha peak values in **Fig. 6 (A-E)**. A significant decrease is found in EO global alpha frequency peak power between pre and post dance conditions (p < 0.05), shown in **Fig. 6A**. **Fig. 6 (C-E)** displays the analysis for EC. ECs are found to have a significant difference (p < 0.05) in both alpha frequency peak power and alpha peak value, in **Fig. 6C** and **Fig. 6D**.

**Figure 6.**
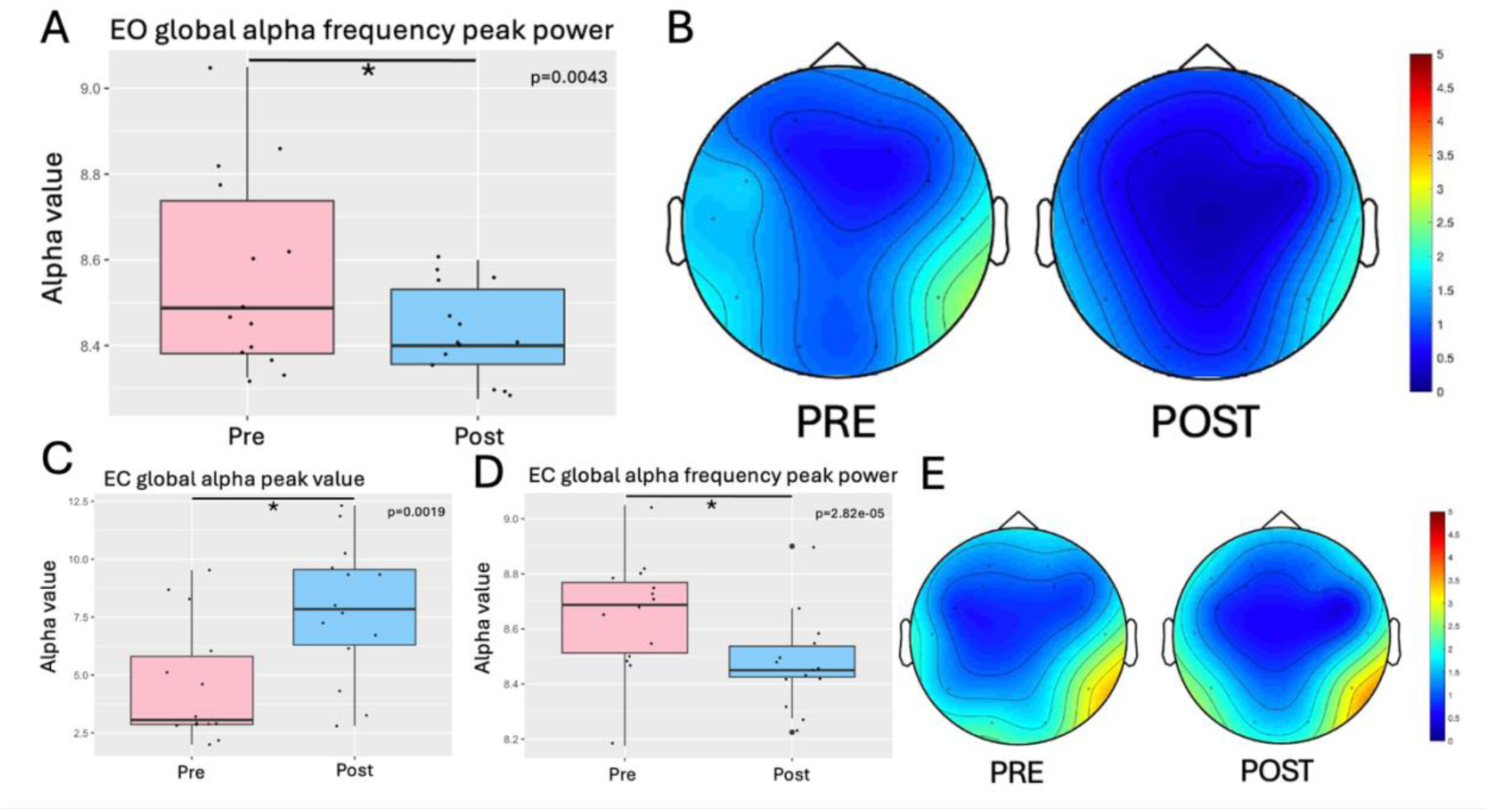
All pre and post rsEEG recording sessions (n=6). **(A)** EO global alpha frequency peak power. X-axis is the pre-dance and post-dance conditions. Each point represents a channel on the EEG headset. Average of each channel was taken for plotting. The y-axis represents the alpha frequency peak power value. All sessions available were represented in this graph. This includes all 6 participants, with an average of 3.33 scans per participant. A significant difference is found between pre- and post-dance conditions (p = 0.004331791). **(B)** EO pre- and post-dance conditions. Scale represents alpha power. **(C)** EC global alpha peak value. X-axis is the pre-dance and post-dance conditions. Each point represents a channel on the EEG headset. Average of each channel was taken for plotting. The Y-axis represents the alpha frequency peak power value. All sessions available were represented in this graph. This includes all 6 participants, with an average of 3.33 EEG scans per participant. There is a significant difference (p = 0.001857791) between pre and post conditions. **(D)** EC global alpha frequency peak power. X-axis is the pre-dance and post-dance conditions. Each point represents a channel on the EEG headset. Average of each channel was taken for plotting. The y-axis represents the alpha frequency peak power value. All sessions available were represented in this graph. This includes all 6 participants, with an average of 3.33 scans per participant. There is a significant difference (p = 2.825375e-05) between pre and post conditions. **(E)** EC pre- and post-dance condition. Scale represents alpha power.

**Fig. 7 (A-C)** performs the same analysis but using the average channel value for participants who recorded more than one session. The across sessions for each channel was calculated and used to plot. Head diagrams could not be represented in this way. Significant differences (p < 0.05) in both EO and EC conditions, as shown in **Fig. 7 (A-C)**. From that average calculation, the frontal cortex (FC) channel values were extracted and plotted (F3, F4, FC5, FC6) in **Fig. 7D** and **Fig. 7E**. A diagram of the EEG cap can be found in the Supplementary materials (S8). T-tests revealed significant differences (p < 0.05) between pre and post dance conditions in the EC condition, but not EO, as shown in **Fig. 7 (D-E)**.

**Figure 7.**
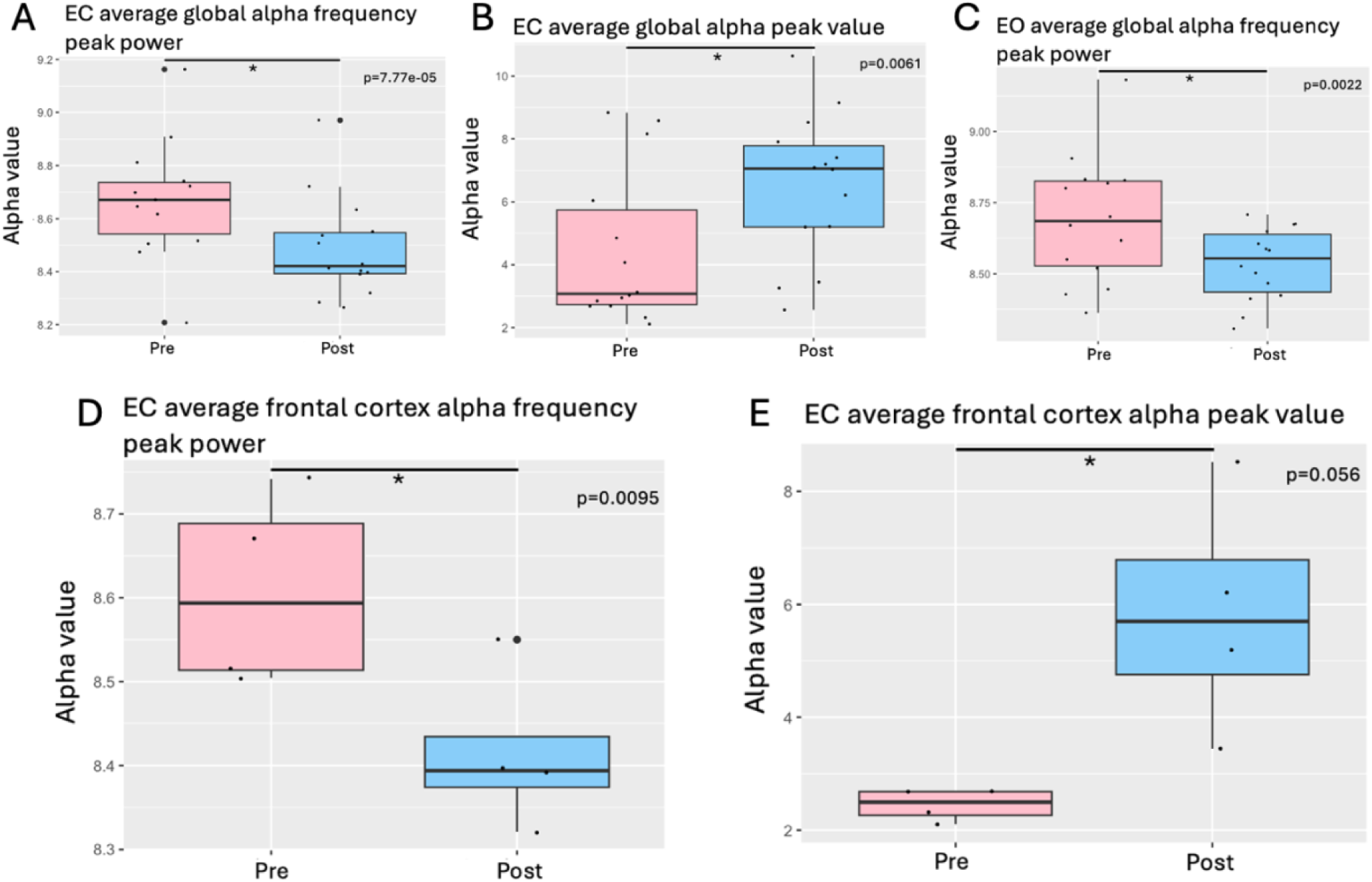
**(A-C)** These graphs represent the average global alpha frequency. For participants with more than one rsEEG session, the average of their sessions was taken and plotted. Each dot represents a channel on the EEG headset. The y-axis represents the numerical value for either the alpha frequency or the alpha peak value. The average EC global alpha frequency peak power shows a significant difference between pre and post conditions (p = 7.768452e-05). The average EC global alpha peak value shows a significant difference between pre and post dance conditions (p = 0.006141888). The average EO global alpha frequency peak power shows a significant difference between pre and post dance conditions (p = 0.002196444). **(D-E)** Four frontal cortex (FC) channels were extracted (F3, F4, FC5, FC6) from the EEG headset and plotted. For participants who had more than one session, the average across sessions was used. Channel values were then averaged across participants and plotted. Each dot represents a FC channel. EC average for FC alpha frequency peak power shows a significant difference between pre and post conditions (p = 0.009495149). EC average for FC alpha peak value shows a significant difference between pre and post conditions (p = 0.05648314).

As shown in **Fig. 8 (A-D),** each participant’s first pre-dance rsEEG recording and last post-dance rsEEG recording was extracted and plotted to show the longitudinal effects of dance. Each participant’s first recording was recorded in 2014, with their last session recorded ranging from 2014 to 2017. A significant increase (p < 0.05) in EO alpha peak values is demonstrated in **Fig. 8A** and **Fig. 8B**. **Fig. 8(C-D)** demonstrate this in EC, with a significant increase (p < 0.05) in alpha peak value depicted in **Fig. 8C**.

**Figure 8.**
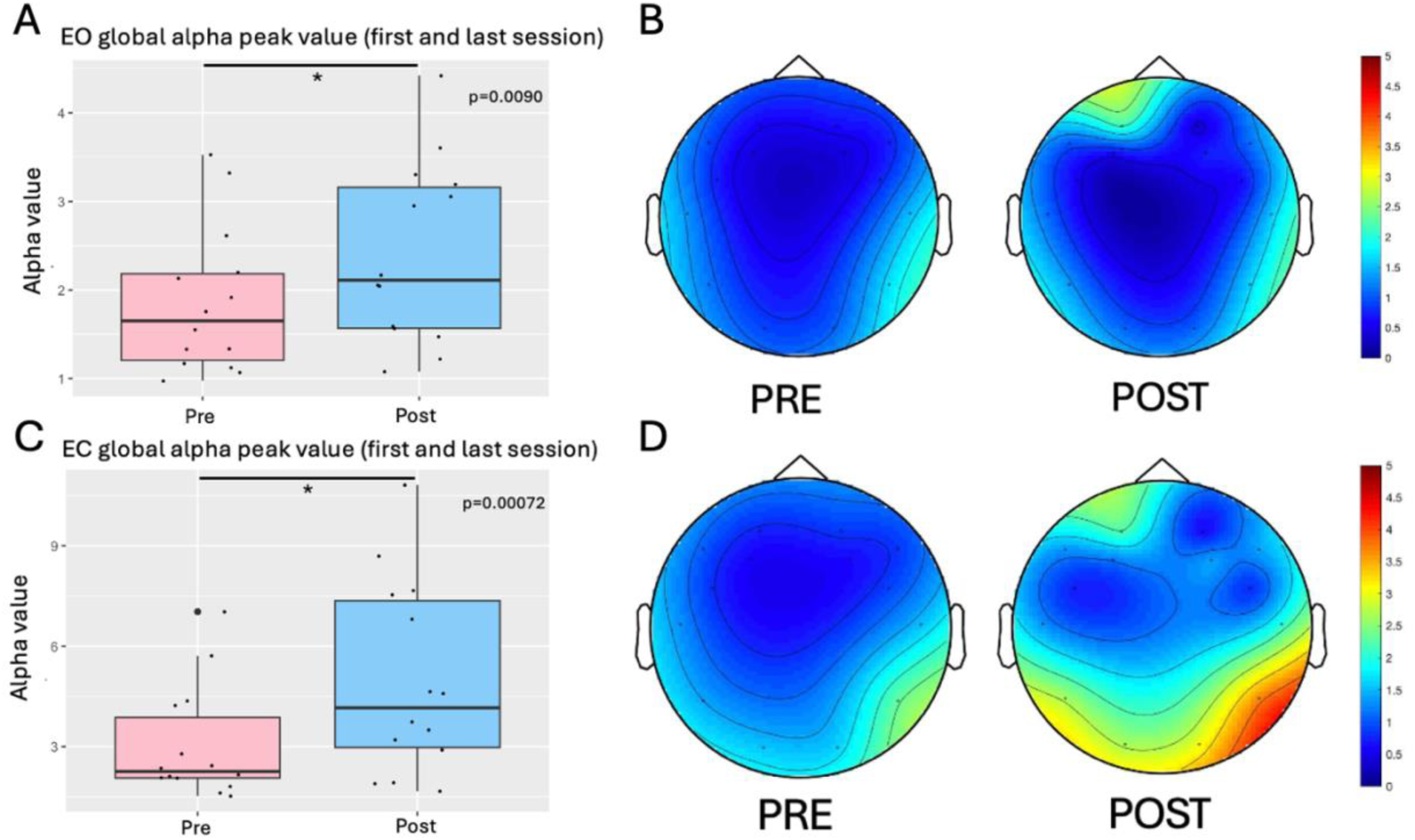
Each participant’s first (pre-dance) and last (post-dance) session were taken and plotted here for EO and EC condition. First scans were all in 2014, and last scans ranged from 2014-2017. **(A)** The average EO global alpha peak value shows a significant difference between pre and post dance conditions (p = 0.009042431). The “Pre” group represents the first pre-dance EO session and the “Post” group represents the last EO post-dance session. Each dot represents a channel in the EEG headset. **(B)** This graph depicts the EO post-dance condition brain wave activity. **(C)** The average EC global alpha peak value shows a significant difference between pre and post dance conditions (p = 0.0007164016). The “pre-sessions-average” group represents the first pre- dance EC session, and the “post-sessions-average” group represents the last EC post-dance session. Each dot represents a channel in the EEG headset. **(D)** This graph depicts the EC post-dance condition alpha wave activity.

## 4. DISCUSSION

Our study shows that in individuals with PD who dance weekly there are long-term (8- month) functional BOLD changes in cortical and subcortical regions, changes in affective state, and short-term (hourly) plastic changes in alpha rhythms as assessed with EEG.

The findings of dance learning related to BOLD changes firstly suggest functional changes in the reward pathway. A change in BOLD signal is reflective of a change in blood-oxygenation level but does not reflect neural activity. The VTA shows a trend, with an increase in BOLD signal from T2 to T3 over the one month of much rehearsal. The VTA is a region that degenerates in PD (Alberico, Cassell & Narayanan, 2015). It is made up of dopaminergic (DA) neurons and GABA neurons, where GABA neurons inhibit the DA neurons (Bouarab, Thompson & Polter, 2019). These changes in BOLD activation may suggest a change in GABA inhibition that may allow an increased excitatory DA output. This is supported by the significant change in BOLD signal in the rNAc and trend in the lNAc. The VTA projects to the NAc and PFC in the reward pathway (Lewis et al., 2021). The NAc has been implicated in DA reward evaluation, anticipation, or prediction of reward (Misaki et al., 2016) and associative learning between motor behaviours and reward (Gale et al., 2014). There is a decrease in BOLD signal from T1 to T3 (i.e. initial learning to after first performance) in the rNAC, and decreases in BOLD signal between T1 and T3, and T1 and T4 in the left NAc. Both regions show a similar trend of decreased BOLD signal over time. This decrease in activation could not only imply a change in excitatory, but also inhibitory activity. Gale et al. (2014) shows reward and reinforcement activity during learning which suggests that the NAc releases GABA to inhibit DA neurons. The VTA receives inhibition from the NAc, and a decrease in inhibition increases the DA neuron firing in the VTA (Bouarab, Thompson & Polter, 2019). With the increase in activation in the VTA, and the decrease in the NAc, these changes may be reflective of a change of GABA output of the NAc (Bouarab, Thompson & Polter, 2019) to the VTA in individuals with PD. The NAc is implicated in neuropsychiatric symptoms of PD (Mavridis 2014a) and listening to rhythms of groove or music has shown to have a reward on the nucleus accumbens, with a combination of motor and reward regions (Foster Vander Elst et al., 2023; Mavridis 2014b). This change demonstrates that dance learning promotes neuroplasticity in the brain through the reward network, in regions related to the degeneration of PD and depression, in individuals with PD which may be neuroprotection.

The OFC plays a role in depression, being key in emotion and reward (Zhang et. al, 2024). The OFC has been implicated in poor decision-making (Kobayakawa, Tsuruya & Kawamura, 2017). Linking these two concepts, the OFC has also been found to be involved in reward-related decision-making (Rolls, Cheng & Feng, 2020). This decrease in BOLD signal activity in the rOFC between T2 and T3, and the lOFC between T1 and T3 may suggest that the participants felt good about their dance, as they knew their dance the best at T3. At T3, participants had been dancing for 5 months. Again, the change in BOLD signal does not indicate whether there is a change in excitatory and inhibitory signal, so a decrease in the OFC does not indicate that depression worsens.

The ACC was found to decrease in activation over time, with a trend between T1 and T4 in the rACC and a significant decrease from T1 to T4 in the lACC. The ACC was found to have hyper-activity in the high-anxiety groups (Criaud et al., 2021), which speculates that the decrease in BOLD signal in the PD participants may correlate with a decrease in anxiety. The ACC may be more implicated in depression than originally anticipated, having networks connected to the VTA (Wei et al., 2018). The change in activation may also be related to a change in depression. The lateralization of significance may be implicated in changes in abstract thinking.

The region in the rPCC is specifically the retrosplenial cortex (RSC) which is involved in spatial and directional orientation, and spatial/episodic memories (Mitchell et al., 2018) with earlier studies showing it to be involved with emotional words (Maddock, 1999). Perhaps, it can be involved in emotional memories, within episodic memories, especially with its connection to the parahippocampal regions (Mitchell et. al, 2018). While visualizing a dance, a significant peak in activation in the RSC is predicted. This could imply that the memory of dance is the most meaningful at that point in time. In this region, there is also a correlation in positive affect scores and BOLD signal change. This implies that there is an emotional component of the RSC, as positive affect feelings increase with BOLD signals.

The PANAS-X shows changes in group and timing conditions that improve GNA scores. A two-way ANOVA shows a significant difference in group and timing, and post-hoc t-tests (Bonferroni-corrected) reveal significant differences (p<0.05). In **Fig. 2A**, t-tests between PD-pre and HC-pre reveal the differences in negative affect scores prior to the dance class, which is important as it reveals that HC-pre initially have a significantly lower score. In both **Fig. 2A** and **Fig. 2B** the difference in PD-pre and PD-post is significant as it shows a decrease in the scores over time for PwPD. It follows the same decrease in GNA scores as a previous study by Ghanai, Barnstaple and DeSouza (2021). This pattern is like the decrease in negative affect score that the controls follow. This implies that dance improves feelings of negative affect in PwPD. A significant difference between HC-post and PD-post reveals that there is still a drastic difference between the two group’s affective state, even after dance.

The PANAS-X shows changes in group, timing and group:timing conditions from a two-way ANOVA that improve GPA scores. Post-hoc t-tests (Bonferroni-corrected) also reveal significant differences between almost all conditions. In **Fig. 3A** and **Fig. 3B** a significant increase in scores between PD-pre and PD-post reveal that dance increases feelings of positive affect over time and that PwPD follow the same trend as the control group. A previous study by Ghanai, Barnstaple & DeSouza (2021) shows the same trend, with GPA scores improving after a dance class, but for online dance classes during the COVID-19 pandemic. A significant difference in both the longitudinal graph is that there is a significant difference between HC-pre and PD-pre and in both the longitudinal and first sessions graph there is a significant difference in scores between HC-post and PD-post, which shows that there is still more that needs to be done to reduce this gap.

The PANAS-X when divided into anxiety negative affect and anxiety positive affect show trends that align with mood changes in PwPD following dance in previous studies (Lewis et al., 2014; Colombo et al., 2022). The two-way ANOVA results in anxiety negative affect supports dance in decreasing feelings of fear, hostility, shyness, and fatigue, which would improve generalized anxiety disorder (GAD). However, with no significant difference between the PD groups when performing t-tests, we cannot confirm that dance improves anxiety in PwPD. In anxiety negative affect, there are differences between PD-pre and HC-pre in the longitudinal graph, which shows that anxious symptoms are present in PwPD more than in controls. In the first session only graphs, there is a significant decrease in scores between PD- post and HC-post, which shows that more needs to be done to have the scores closer together. In anxiety positive affect, the two-way ANOVA results show significant changes across all conditions, which demonstrates that dance improves attentiveness and serenity, which are affected in those diagnosed with GAD. The same trend is also observed with significant differences between PD-pre and HC-pre in the longitudinal graph, along with significant differences between PD-post and HC-post in both the longitudinal and first session analyses. This demonstrates that more needs to be done to close this gap yet reinforces the concept that PwPD are more affected by these symptoms.

When the PANAS-X is divided into depression negative affect, the two-way ANOVA reveals significant changes across group and timing conditions. The post-hoc t-tests (Bonferroni-corrected) reveal significant differences between PD-pre and PD-post groups in both the longitudinal and first session analyses demonstrating that dance improves sadness, guilt, and fatigue which are affected in those diagnosed with major depressive disorder (MDD). Although this has not previously been done with PANAS-X questionnaires, dance has been shown to improve depression using the Self-rating Depression Scale (SDS) (Hashimoto et al., 2015) and the GDS (Barnstaple et al., 2022). The significantly lower scores from HC-pre to PD-pre display the prevalence of MDD symptoms in PwPD prior to dance. The significant difference even after dance between HC-post and PD-post in the longitudinal and first session analyses further show that although dance improves the symptoms, PwPD are more severely affected by MDD symptoms. With the depression positive affect, the two-way ANOVA reveals significant changes across all conditions in both the longitudinal and first session analyses. An increase in scores between PD-pre and PD-post in both analyses reveals that dance increases feelings of joviality, self-assurance, and attentiveness in MDD. This also follows the increase in score that the controls display. The significant difference between PD-pre and HC-pre in the longitudinal analysis displays the lack of joviality, self-assurance, and attentiveness in PwPD prior to dance, and emphasizes the fact that NMS need to be addressed in PD. These results follow the expected patterns, as the scores improve after dance.

A trend is seen between GPA and change in BOLD signal in the rPCC. This trend shows that as BOLD % signal change increases, PANAS-X average scores increase as well. This shows that there is a connection between the RSC and emotional state. Participants were instructed to visualize the dance in the scanner, which supports the RSC in spatial navigation (Vann, Aggleton & Maguire, 2009; Mitchell et al., 2018), but with the correlation with GPA scores, this questions previous theories on RSC’s involvement in episodic and autobiographical memory and its connection to the limbic system (Chrastil, 2018). As participants are retrieving the memory of them dancing, this would spark positive emotions, which makes this region interesting to continue studying in PwPD.

Studies on rsEEG in PwPD are scarce. An article by Babiloni et al. (2024) found lower alpha activation from EC to EO conditions in individuals with PD dementia (PDD), suggesting desynchronized rsEEG alpha rhythms in posterior cortical regions. Another study by Han et al. (2013) found EEG abnormalities in early PD, with reduced alpha band power. For all participant’s scans, a significant decrease is found in global alpha frequency peak power. This is consistent with previous research (Han et al., 2013; Babiloni et al., 2024) however, with a dance intervention, a different trend than typical research outcomes would be expected. From the diagram, there is a reduction in posterior activity, consistent with previous research. In the EC condition, there is a decrease in global alpha frequency peak power, but an increase in alpha peak value. This significant change implies that dance promotes increased alpha peak values. The diagrams show a change in activation in the left occipital region, along with increased alpha power in the frontal cortex. Participants were not told to visualize anything specific, but it is possible that after dance, they would think of more positive things which would spark this change in activation.

When using the average alpha frequency peak power and alpha peak value for participants who had more than one session, the same trend is seen, where there is a significant decrease in alpha frequency peak power and an increase in alpha peak values in the EC conditions. In EO, a decrease in alpha frequency peak power is only seen. Again, this decrease may be due to the nature of the disease, but the increase in alpha peak value is indicative of a change in the typical PD patterns. Altered alpha oscillations at rest are found in PD (Ye et al., 2022), therefore any changes found due to an intervention are impactful.

In isolating the four FC channels (F3, F4, FC5, FC6) significant changes were only observed in the EC condition. It appears that when there is a decrease in alpha frequency peak power, there are increases in the alpha peak value. This indicates that dance induces changes in the FC related to alpha oscillations.

Participants’ first and last rsEEG sessions were taken and plotted to show the longitudinal impact of dance on the participant. A significant increase in global alpha peak value was found in the EO condition, with an increase in alpha oscillations in the medial left FC and in the bilateral parietal cortex. With the frontal lobe being involved in emotional processing, this result may reflect the social nature of dancing, and the emotions it produces. There is a dramatic increase in alpha oscillations in the EC condition, with an increase in alpha peak values. In the diagram, there is a global increase in activation, particularly in the posterior regions of the brain (occipital lobe, parietal lobes). With EC, participants can be visualizing anything, or nothing, which many account for this dramatic change. However, this shows the longitudinal effects of dance, in that the longer the participants dance, the more alpha power peak values increase. With the small body of research on EEG in PwPD, any significant change in alpha power is relevant, and supports dance as a form of neurorehabilitation affecting alpha oscillations.

No significant correlations were found between rsEEG alpha peak values and changes in PANAS-X scores. This may be due to the nature of the rsEEG recordings, as they were not instructed to think about the dance itself. The participants thinking randomly would not evoke emotional states or activate these neural circuits. Additionally, EEG only records cortical levels, whereas many brain regions involved in the reward pathway and the limbic system are subcortical (e.g. VTA, NAc, etc.). In addition, we only had six subjects in this report that had both fMRI and EEG and we need to expand on this in future studies. But recall this was a community dance class with the fMRI over 8 months and EEG over 6 years in individuals with Parkinson’s disease and their caregivers dancing and acting as controls in our study.

One limitation of the study is that it does not include PwPD or controls who do not partake in the dance classes. This would enable the evaluation of the effectiveness of dance on PwPD and controls. Additionally, there is no overlap between the rsEEG, fMRI and PANAS-X. Ideally, having a study that better utilizes these methods would aid in attempting to correlate the multimodal recordings. Without the rsEEG evoking emotional states, it is challenging to interpret the results. An evoked response study that has conditions on EEG such as visualizing the dance would be beneficial as it would reflect the fMRI protocol and better understand the underlying circuitry. If this study were replicated, the use of anxiety and depression-based surveys with more support as evaluation methods of GAD and MDD might enhance the study’s brain behaviour relationships.

Overall, this study supports dance as a form of neurorehabilitation. This study shows that dance improves NMS of anxiety and depression, along with GNA and GPA. These improvements in scores are supported by the neuroplasticity observed in the reward pathway (i.e., VTA, NAc, and OFC), the ACC and PCC, along with the changes in alpha oscillations. Future research should be conducted to support this conclusion in a greater number of subjects using a randomized controlled design across multi sites.

## Data Availability

All data produced in the present study are available upon reasonable request to the authors

## FUNDING

This work was supported in part by a Parkinson Canada Pilot Grant awarded to Joseph F.X. DeSouza. Joseph F.X. DeSouza is also funded by a Natural Sciences and Engineering Research Council (NSERC) Discovery Grant (2017-05647) and generous donations from the Irpinia Club of Toronto and other supporters.

## ETHICAL CONSIDERATIONS

Written informed consent was obtained at each data collection time point throughout the 8- month data collection period, following an approved protocol from York University’s Ethics Board (2013-211 & 2017-296). The most recent Ethics approval (Certificate #: 2025-032) was obtained on February 21, 2025, from the York University Office of Research Ethics. All procedures adhered to the ethical standards of the institutional and national committees on human experimentation and complied with the Helsinki Declaration of 1975, as revised in 2000. Participants’ privacy and confidentiality were ensured through de-identification, and data were securely stored on password-protected external hard drives in the research laboratory at York University. Participants received CAD $25 per imaging session and CAD $50 reimbursement per visit to York University for imaging-related travel costs (CAD $1 ≈ USD $0.71).

## DATA AVAILABILITY STATEMENT

The study data is available from the corresponding author on reasonable request.

## DECLARATION OF COMPETING INTEREST

The authors declare that they have no conflict of interest.

## ACKNOWLEDGMENTS

We extend our sincere gratitude to all members of our laboratory over the years who contributed to this project, as well as to the individuals at our various testing locations for their support. We are indebted to S. Robichaud, D. Rabinovich, R. Cohan, P. Dhami, S. Maguire, H. Tehrani, K. McDonald, and R. Andrew for invaluable assistance during all phases of the research program. We are especially grateful for the collaborative efforts and continued support from Canada’s National Ballet School (A. Seto and A. Powell), Dance with Parkinson’s Canada (S. Robichaud), Trinity St. Paul’s Church in Toronto, and the Dance for Parkinson’s program at the Mark Morris Dance Group (D. Leventhal).

## S. SUPPLEMENTARY MATERIAL

**Table S1.** Talairach coordinates of ROIs presented in results.

| ROI | Hemisphere | Talairach Coordinates |  |  | Number of voxels |
| --- | --- | --- | --- | --- | --- |
|  |  | *x* | *y* | *z* |  |
| VTA |  | -2 | -23 | -13 | 515 |
| NAc | Right | 11 | 9 | -6 | 515 |
|  | Left | -12 | 9 | -6 | 515 |
| OFC | Right | 11 | 47 | -15 | 515 |
|  | Left | -10 | 41 | -15 | 925 |
| ACC | Right | 4 | 30 | 24 | 925 |
|  | Left | -9 | 26 | 27 | 925 |
| PCC | Right | -3 | -39 | 11 | 515 |

**Table S2.**
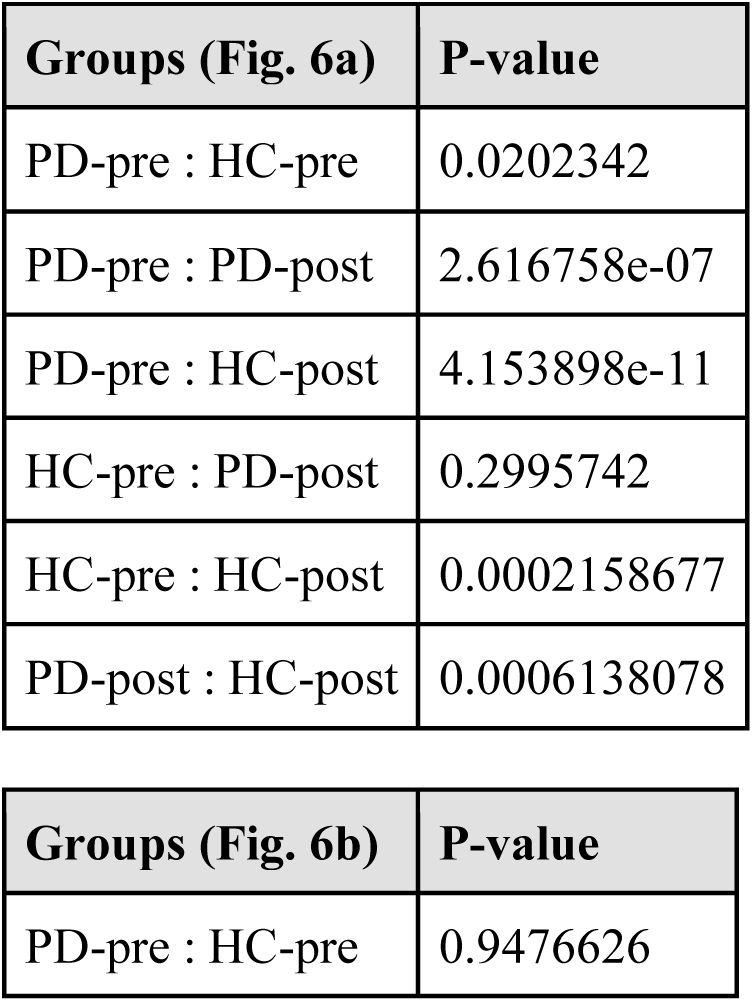
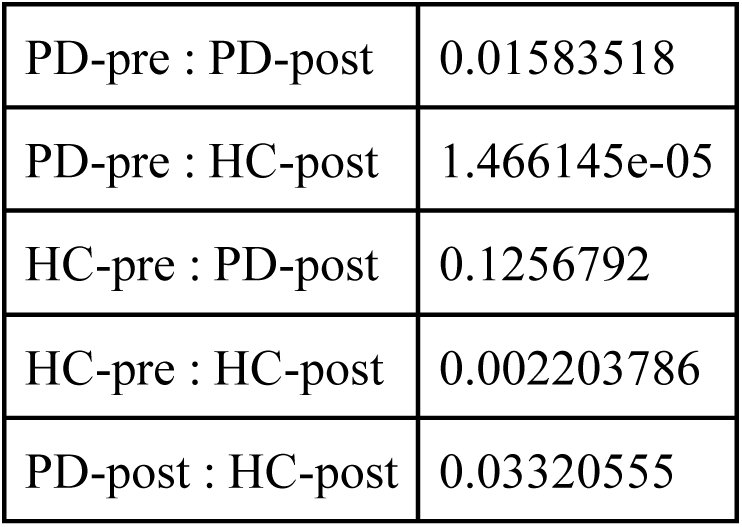
P-values of post-hoc t-tests (Bonferroni-corrected) for PANAS-X scores.

| Groups (Fig. 6a) | P-value |
| --- | --- |
| PD-pre : HC-pre | 0.0202342 |
| PD-pre : PD-post | 2.616758e-07 |
| PD-pre : HC-post | 4.153898e-11 |
| HC-pre : PD-post | 0.2995742 |
| HC-pre : HC-post | 0.0002158677 |
| PD-post : HC-post | 0.0006138078 |

| Groups (Fig. 6b) | P-value |
| --- | --- |
| PD-pre : HC-pre | 0.9476626 |
| PD-pre : PD-post | 0.01583518 |
| PD-pre : HC-post | 1.466145e-05 |
| HC-pre : PD-post | 0.1256792 |
| HC-pre : HC-post | 0.002203786 |
| PD-post : HC-post | 0.03320555 |

**Table S3.** P-values of post-hoc t-tests (Bonferroni-corrected) for PANAS-X scores.

| Groups | P-value |
| --- | --- |
| PD-pre : HC-pre | 0.001739216 |
| PD-pre : PD-post | 0.0009047735 |
| PD-pre : HC-post | 2.390414e-10 |
| HC-pre : PD-post | 0.06806415 |
| HC-pre : HC-post | 5.745924e-05 |
| PD-post : HC-post | 3.644058e-07 |
| PD-pre : HC-pre | 0.1284591 |
| PD-pre : PD-post | 0.05938672 |
| PD-pre : HC-post | 3.646829e-05 |
| HC-pre : PD-post | 0.5028496 |
| HC-pre : HC-post | 0.001200797 |
| PD-post : HC-post | 0.0006558629 |

**Table S4.** P-values of post-hoc t-tests (Bonferroni-corrected) for PANAS-X scores.

| Groups | P-value |
| --- | --- |
| PD-pre : HC-pre | 0.005070116 |
| PD-pre : PD-post | 0.1382452 |
| PD-pre : HC-post | 1.833435e-07 |
| HC-pre : PD-post | 0.08425504 |
| HC-pre : HC-post | 0.03138455 |
| PD-post : HC-post | 7.631443e-05 |
| PD-pre : HC-pre | 0.5783268 |
| PD-pre : PD-post | 0.7555587 |
| PD-pre : HC-post | 0.02466926 |
| HC-pre : PD-post | 0.4754941 |
| HC-pre : HC-post | 0.1701372 |
| PD-post : HC-post | 0.02736014 |

**Table S5.** P-values of post-hoc t-tests (Bonferroni-corrected) for PANAS-X scores.

| Groups | P-value |
| --- | --- |
| PD-pre : HC-pre | 0.01955731 |
| PD-pre : PD-post | 0.2948346 |
| PD-pre : HC-post | 2.305384e-07 |
| HC-pre : PD-post | 0.05940018 |
| HC-pre : HC-post | 6.579055e-05 |
| PD-post : HC-post | 1.390648e-06 |
| PD-pre : HC-pre | 0.3823305 |
| PD-pre : PD-post | 0.3026728 |
| PD-pre : HC-post | 0.001192465 |
| HC-pre : PD-post | 0.6777087 |
| HC-pre : HC-post | 0.0007818618 |
| PD-post : HC-post | 0.003781294 |

**Table S6.** P-values of post-hoc t-tests (Bonferroni-corrected) for PANAS-X scores.

| Groups | P-value |
| --- | --- |
| PD-pre : HC-pre | 0.03215561 |
| PD-pre : PD-post | 5.712204e-10 |
| PD-pre : HC-post | 1.519137e-15 |
| HC-pre : PD-post | 0.02987222 |
| HC-pre : HC-post | 2.954306e-07 |
| PD-post : HC-post | 0.0006659864 |
| PD-pre : HC-pre | 0.9821897 |
| PD-pre : PD-post | 0.001184357 |
| PD-pre : HC-post | 3.032919e-06 |
| HC-pre : PD-post | 0.02993754 |
| HC-pre : HC-post | 0.0002954272 |
| PD-post : HC-post | 0.03864952 |

**Table S7.** P-values of post-hoc t-tests (Bonferroni-corrected) for PANAS-X scores.

| Groups | P-value |
| --- | --- |
| PD-pre : HC-pre | 0.004122958 |
| PD-pre : PD-post | 0.0002924221 |
| PD-pre : HC-post | 3.473775e-09 |
| HC-pre : PD-post | 0.1388551 |
| HC-pre : HC-post | 4.992404e-05 |
| PD-post : HC-post | 4.513376e-06 |
| PD-pre : HC-pre | 0.2368239 |
| PD-pre : PD-post | 0.04343472 |
| PD-pre : HC-post | 0.0005091737 |
| HC-pre : PD-post | 0.7794253 |
| HC-pre : HC-post | 0.001395617 |
| PD-post : HC-post | 0.007195424 |

## Notes

### Competing Interest Statement

The authors have declared no competing interest.

### Funding Statement

This study did not receive any funding

### Author Declarations

Ethics committee/IRB of York University gave ethical approval for this work. (Certificate numbers: 2013-211, 2017-296, and 2025-032)

## REFERENCES

Alberico, S. L., Cassell, M. D., & Narayanan, N. S. (2015). The vulnerable ventral tegmental area in parkinson’s disease. Basal Ganglia, 5(2–3), 51–55. 10.1016/j.baga.2015.06.001

Allen, M., Fardo, F., Dietz, M. J., Hillebrandt, H., Friston, K. J., Rees, G., & Roepstorff, A. (2016). Anterior insula coordinates hierarchical processing of tactile mismatch responses. NeuroImage, 127, 34–43. 10.1016/j.neuroimage.2015.11.030

Asmundson, G. J., Fetzner, M. G., DeBoer, L. B., Powers, M. B., Otto, M. W., & Smits, J. A. (2013). Let’s get physical: A contemporary review of the anxiolytic effects of exercise for anxiety and its disorders. Depression and Anxiety, 30(4), 362–373. 10.1002/da.22043

Babiloni, C., Noce, G., Tucci, F., Jakhar, D., Ferri, R., Panerai, S., Catania, V., Soricelli, A., Salvatore, M., Nobili, F., Arnaldi, D., Famà, F., Buttinelli, C., Giubilei, F., Onofrj, M., Stocchi, F., Vacca, L., Radicati, F., Fuhr, P., … Del Percio, C. (2024). Poor reactivity of posterior electroencephalographic alpha rhythms during the eyes open condition in patients with dementia due to parkinson’s disease. Neurobiology of Aging, 135, 1–14. 10.1016/j.neurobiolaging.2023.11.010

Barnstaple, R. E., Bearss, K., Bar, R. J., & DeSouza, J. F. (2022). Weekly Dance Training over Eight Months Reduces Depression and Correlates with fMRI Brain Signals in Subcallosal Cingulate Gyrus (SCG) for People with Parkinson’s Disease: An Observational Study. BioRxiv 10.1101/2022.10.14.512180

Barnstaple, R. E., & DeSouza, J. F. X. (2018). Dance and neurorehabilitation - mixed-methods research models. *Functional Neurology*, Rehabilitation and Ergonomics, 7(1).

Bearss, K. A., & DeSouza, J. F. X. (2021). Parkinson’s disease motor symptom progression slowed with multisensory dance learning over 3-years: A preliminary longitudinal investigation. Brain Sciences, 11(7), 895. 10.3390/brainsci11070895

Bouarab, C., Thompson, B., & Polter, A. M. (2019). VTA GABA neurons at the interface of stress and reward. Frontiers in Neural Circuits, 13. 10.3389/fncir.2019.00078

Broen, M. P., Narayen, N. E., Kuijf, M. L., Dissanayaka, N. N., & Leentjens, A. F. (2016). Prevalence of anxiety in parkinson’s disease: A systematic review and meta-analysis. Movement Disorders, 31(8), 1125–1133. 10.1002/mds.26643

Carey, G., Görmezoğlu, M., Jong, J. J. A., Hofman, P. A. M., Backes, W. H., Dujardin, K., & Leentjens, A. F. G. (2020). Neuroimaging of Anxiety in Parkinson’s Disease: A Systematic Review. Movement Disorders, 36(2), 327–339. 10.1002/mds.28404

Chrastil, E. R. (2018). Heterogeneity in human retrosplenial cortex: A review of function and connectivity. Behavioral Neuroscience, 132(5), 317–338. 10.1037/bne0000261

Criaud, M., Kim, J.-H., Zurowski, M., Lobaugh, N., Chavez, S., Houle, S., & Strafella, A. P. (2021). Anxiety in parkinson’s disease: Abnormal resting activity and connectivity. Brain Research, 1753, 147235. 10.1016/j.brainres.2020.147235

Colombo, B., Rigby, A., Gnerre, M., & Biassoni, F. (2022). The effects of a dance and music-based intervention on parkinson’s patients’ well-being: An interview study. International Journal of Environmental Research and Public Health, 19(12), 7519. 10.3390/ijerph19127519

DeSouza, J. F., Simon, J. R., Karimi, A., Agrawal, A., Barnstaple, R. E., Bek, J., Bar, R., Bearss, K., & Ghanai, K. (2023). Modulations of sensorimotor network through visual motor training in people with parkinson’s disease (PwPD). Journal of Vision, 23(9), 5423. 10.1167/jov.23.9.5423

Ding, Y.-D., Chen, X., Chen, Z.-B., Li, L., Li, X.-Y., Castellanos, F. X., Bai, T.-J., Bo, Q.-J., Cao, J., Chang, Z.-K., Chen, G.-M., Chen, N.-X., Chen, W., Cheng, C., Cheng, Y.-Q., Cui, X.-L., Duan, J., Fang, Y.-R., Gong, Q.-Y., … Guo, W.-B. (2022). Reduced nucleus accumbens functional connectivity in reward network and default mode network in patients with recurrent major depressive disorder. Translational Psychiatry, 12(1). 10.1038/s41398-022-01995-x

Di Nota, P. M., Chartrand, J. M., Levkov, G. R., Montefusco-Siegmund, R., & DeSouza, J. F. X. (2017). Experience-dependent modulation of alpha and beta during action observation and motor imagery. BMC neuroscience, 18(1), 28. 10.1186/s12868-017-0349-0

Fontanesi, C., & DeSouza, J. F. (2021). Beauty that moves: Dance for parkinson’s effects on affect, self-efficacy, gait symmetry, and dual task performance. Frontiers in Psychology, 11. 10.3389/fpsyg.2020.600440

Foster Vander Elst, O., Foster, N. H. D., Vuust, P., Keller, P. E., & Kringelbach, M. L. (2023). The Neuroscience of Dance: A Conceptual Framework and systematic review. Neuroscience & Biobehavioral Reviews, 150, 105197. 10.1016/j.neubiorev.2023.105197

Gale, J. T., Shields, D. C., Ishizawa, Y., & Eskandar, E. N. (2014). Reward and reinforcement activity in the nucleus accumbens during learning. Frontiers in Behavioral Neuroscience, 8. 10.3389/fnbeh.2014.00114

Garlovsky, J. K., Overton, P. G., & Simpson, J. (2016). Psychological predictors of anxiety and depression in parkinson’s disease: A systematic review. Journal of Clinical Psychology, 72(10), 979–998. 10.1002/jclp.22308

Ghanai, K., Barnstaple, R. E., & DeSouza, J. F. (2021). Virtually in Synch: A pilot study on affective dimensions of dancing with parkinson’s during COVID-19. Research in Dance Education, 1–15. 10.1080/14647893.2021.2005560

Gong, L., Xu, R., Yang, D., Wang, J., Ding, X., Zhang, B., Zhang, X., Hu, Z., & Xi, C. (2022). Orbitofrontal cortex functional connectivity-based classification for chronic insomnia disorder patients with depression symptoms. Frontiers in Psychiatry, 13. 10.3389/fpsyt.2022.907978

Gutmann, B., Mierau, A., Hülsdünker, T., Hildebrand, C., Przyklenk, A., Hollmann, W., & Strüder, H. K. (2015). Effects of physical exercise on individual resting state EEG alpha peak frequency. Neural Plasticity, 2015, 1–6. 10.1155/2015/717312

Han, C.-X., Wang, J., Yi, G.-S., & Che, Y.-Q. (2013). Investigation of EEG abnormalities in the early stage of parkinson’s disease. Cognitive Neurodynamics, 7(4), 351–359. 10.1007/s11571-013-9247-z

Hashimoto, H., Takabatake, S., Miyaguchi, H., Nakanishi, H., & Naitou, Y. (2015). Effects of dance on motor functions, cognitive functions, and mental symptoms of parkinson’s disease: A quasi-randomized pilot trial. Complementary Therapies in Medicine, 23(2), 210–219. 10.1016/j.ctim.2015.01.010

Houston, S., & McGill, A. (2013). A mixed-methods study into ballet for people living with Parkinson’s. Arts & Health, 5(2), 103–119. 10.1080/17533015.2012.745580

Kobayakawa, M., Tsuruya, N., & Kawamura, M. (2017). Decision-making performance in parkinson’s disease correlates with lateral orbitofrontal volume. Journal of the Neurological Sciences, 372, 232–238. 10.1016/j.jns.2016.11.046

Kshtriya, S., Barnstaple, R., Rabinovich, D. B., & DeSouza, J. F. (2015). Dance and aging: A critical review of findings in Neuroscience. American Journal of Dance Therapy, 37(2), 81–112. 10.1007/s10465-015-9196-7

Lewis, C., Annett, L. E., Davenport, S., Hall, A. A., & Lovatt, P. (2014). Mood changes following social dance sessions in people with parkinson’s disease. Journal of Health Psychology, 21(4), 483–492. 10.1177/1359105314529681

Lewis, R. G., Florio, E., Punzo, D., & Borrelli, E. (2021). The Brain’s reward system in health and disease. Circadian Clock in Brain Health and Disease, 57–69. 10.1007/978-3-030-81147-1_4

Maddock, R. J. (1999). The retrosplenial cortex and emotion: New insights from functional neuroimaging of the human brain. Trends in Neurosciences, 22(7), 310–316. 10.1016/s0166-2236(98)01374-5

Mamiya, P. C., Richards, T., Corrigan, N. M., & Kuhl, P. K. (2020). Strength of ventral tegmental area connections with left caudate nucleus is related to conflict monitoring. Frontiers in Psychology, 10. 10.3389/fpsyg.2019.02869

Mavridis, I. N. (2014a). Is nucleus accumbens atrophy correlated with cognitive symptoms of parkinson’s disease? Brain, 138(1). 10.1093/brain/awu197

Mavridis, I. N. (2014b). Music and the nucleus accumbens. Surgical and Radiologic Anatomy, 37(2), 121–125. 10.1007/s00276-014-1360-0

Miller, D. B., & O’Callaghan, J. P. (2015). Biomarkers of parkinson’s disease: Present and future. Metabolism, 64(3). 10.1016/j.metabol.2014.10.030

Misaki, M., Suzuki, H., Savitz, J., Drevets, W. C., & Bodurka, J. (2016). Individual variations in nucleus accumbens responses associated with major depressive disorder symptoms. Scientific Reports, 6(1). 10.1038/srep21227

Mitchell, A. S., Czajkowski, R., Zhang, N., Jeffery, K., & Nelson, A. J. (2018). Retrosplenial cortex and its role in spatial cognition. Brain and Neuroscience Advances, 2, 239821281875709. 10.1177/2398212818757098

Mochcovitch, M. D., da Rocha Freire, R. C., Garcia, R. F., & Nardi, A. E. (2014). A systematic review of fmri studies in generalized anxiety disorder: Evaluating its neural and cognitive basis. Journal of Affective Disorders, 167, 336–342. 10.1016/j.jad.2014.06.041

Olde Dubbelink, K. T. E., Stoffers, D., Deijen, J. B., Twisk, J. W. R., Stam, C. J., & Berendse, H. W. (2013). Cognitive decline in parkinson’s disease is associated with slowing of resting-state brain activity: A longitudinal study. Neurobiology of Aging, 34(2), 408–418. 10.1016/j.neurobiolaging.2012.02.029

Peterson, A. C., Zhang, S., Hu, S., Chao, H. H., & Li, C. R. (2017). The effects of age, from young to middle adulthood, and gender on resting state functional connectivity of the dopaminergic midbrain. Frontiers in Human Neuroscience, 11. 10.3389/fnhum.2017.00052

Rabinovich, D. B., Garretto, N. S., Arakaki, T., & DeSouza, J. F. (2021). A high dose tango intervention for people with parkinson’s disease (PwPD). Advances in Integrative Medicine, 8(4), 272–277. 10.1016/j.aimed.2021.07.005

Reijnders, J. S. A. M., Ehrt, U., Weber, W. E. J., Aarsland, D., & Leentjens, A. F. G. (2007). A systematic review of prevalence studies of depression in parkinson’s disease. Movement Disorders, 23(2), 183–189. 10.1002/mds.21803

Rolls, E. T., Cheng, W., & Feng, J. (2020). The orbitofrontal cortex: Reward, emotion and Depression. Brain Communications, 2(2). 10.1093/braincomms/fcaa196

Simon, J. R., Bek, J., Ghanai, K., Bearss, K., Barnstaple, R. E., Bar, R. J., & DeSouza, J. F. (2023). Neural Effects of Multisensory Dance Training in Parkinson’s Disease: A Longitudinal Neuroimaging Case Study. 10.31234/osf.io/kwy9x

Stewart, S. A. (2005). The effects of benzodiazepines on cognition. Journal of Clinical Psychiatry, 66(2), 9–13.

Stoker, T. B., & Barker, R. A. (2020). Recent developments in the treatment of parkinson’s disease. F1000Research, 9, 862. 10.12688/f1000research.25634.1

Su, D., Cui, Y., Liu, Z., Chen, H., Fang, J., Ma, H., Zhou, J., & Feng, T. (2022). Altered Brain Activity in Depression of Parkinson’s Disease: A Meta-Analysis and Validation Study. Frontiers in Aging Neuroscience, 14. 10.3389/fnagi.2022.806054

Sundermann, B., Olde lütke Beverborg, M., & Pfleiderer, B. (2014). Toward literature-based feature selection for diagnostic classification: A meta-analysis of resting-state fmri in depression. Frontiers in Human Neuroscience, 8. 10.3389/fnhum.2014.00692

Vann, S. D., Aggleton, J. P., & Maguire, E. A. (2009). What does the retrosplenial cortex do? Nature Reviews Neuroscience, 10(11), 792–802. 10.1038/nrn2733

Watson, D., & Clark, L. A. (1994). The panas-X: Manual for the positive and negative affect schedule - expanded form. University of Iowa. 10.17077/48vt-m4t2

Wei, L., Hu, X., Yuan, Y., Liu, W., & Chen, H. (2018). Abnormal ventral tegmental area-anterior cingulate cortex connectivity in Parkinson’s disease with depression. Behavioural Brain Research, 347, 132–139. 10.1016/j.bbr.2018.03.011

Westheimer, O., McRae, C., Henchcliffe, C., Fesharaki, A., Glazman, S., Ene, H., & Bodis-Wollner, I. (2015). Dance for PD: A preliminary investigation of effects on motor function and quality of life among persons with parkinson’s disease (PD). Journal of Neural Transmission, 122(9), 1263–1270. 10.1007/s00702-015-1380-x

Ye, Z., Heldmann, M., Herrmann, L., Brüggemann, N., & Münte, T. F. (2022). Altered alpha and theta oscillations correlate with sequential working memory in parkinson’s disease. Brain Communications, 4(3). 10.1093/braincomms/fcac096

Yi, G.-S., Wang, J., Deng, B., & Wei, X.-L. (2016). Complexity of resting-state EEG activity in the patients with early-stage parkinson’s disease. Cognitive Neurodynamics, 11(2), 147–160. 10.1007/s11571-016-9415-z

Zhang, B., Rolls, E. T., Wang, X., Xie, C., Cheng, W., & Feng, J. (2024). Roles of the medial and lateral orbitofrontal cortex in major depression and its treatment. Molecular Psychiatry. 10.1038/s41380-023-02380-w

Zhao, X.-H., Wang, P.-J., Li, C.-B., Hu, Z.-H., Xi, Q., Wu, W.-Y., & Tang, X.-W. (2007). Altered default mode network activity in patient with anxiety disorders: An fmri study. European Journal of Radiology, 63(3), 373–378. 10.1016/j.ejrad.2007.02.006

Zhao, X.H., Wang, P.J., Li, C., Wang, J.H., Yang, Z.Y., Hu, Z.H., & Wu, W.Y. (2006). [Prefrontal and superior temporal lobe hyperactivity as a biological substrate of generalized anxiety disorders]. Europe PMC, 86(14), 955–960.

